# What have we learned about positive changes experienced during COVID-19 lockdown? Evidence of the social patterning of change

**DOI:** 10.1101/2020.09.24.20200865

**Authors:** Lynn Williams, Lesley Rollins, David Young, Leanne Fleming, Madeleine Grealy, Xanne Janssen, Alison Kirk, Bradley MacDonald, Paul Flowers

## Abstract

**Background:** Multiple studies have highlighted the negative impact of COVID-19 and its particular effects on vulnerable sub-populations. Complementing this work, here, we report on the social patterning of self-reported positive changes experienced during COVID-19 national lockdown in Scotland.

**Methods:** The CATALYST study collected data from 3342 adults in Scotland during weeks 9-12 of a national lockdown. Using a cross-sectional design, participants completed an online questionnaire providing data on key sociodemographic and health variables, and completed a measure of positive change. The positive change measure spanned diverse domains (e.g., more quality time with family, developing new hobbies, more physical activity, and better quality of sleep). We used univariate analysis and stepwise regression to examine the contribution of a range of sociodemographic factors (e.g., age, gender, ethnicity, educational attainment, and employment status) in explaining positive change.

**Results:** There were clear sociodemographic differences across positive change scores. Those reporting higher levels of positive change were female, from younger age groups, married or living with their partner, employed, and in better health.

**Conclusion:** Overall our results highlight the social patterning of positive changes during lockdown in Scotland. These findings begin to illuminate the complexity of the unanticipated effects of national lockdown and will be used to support future intervention development work sharing lessons learned from lockdown to increase positive health change amongst those who may benefit.

## Introduction

In many countries, COVID-19 national lockdowns have been the most profound, deep reaching, and significant public health interventions within living memory. Fig 1 provides a logic model describing a high-level overview of lockdown as a complex public health intervention. It shows a range of key contextual elements important to understanding the situation in which lockdown has taken place and it shows the central problem that initial lockdown was intended to resolve (i.e. exponential transmission of COVID-19). It also highlights the complexity of lockdown as a public health intervention, with multiple, interdependent components, cumulatively working through varied and intersecting mechanisms to elicit a range of intended and unintended positive and negative changes.

**Fig 1.**
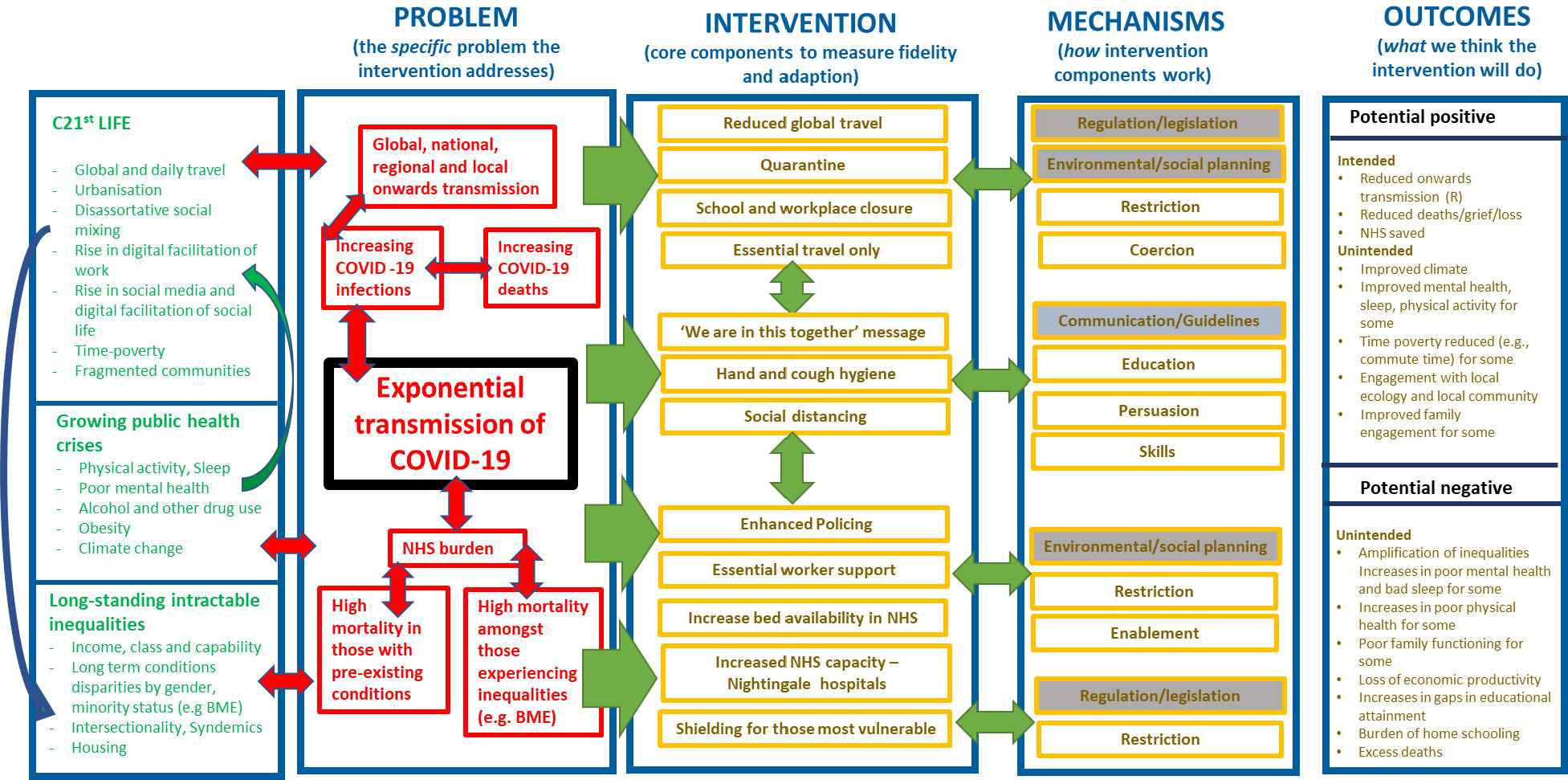
Theorising the initial effects of national ‘lockdown’ for COVID-19: Logic model showing high-level overview.

Within Scotland it is clear that the initial national lockdown intervention succeeded in relation to its primary goal of reducing the exponential transmission of COVID-19 and achieving its intended positive health outcomes (see Fig 1). Similarly, as had been anticipated, lockdowns have led to unintended and negative health consequences. For example, there is emerging evidence concerning the amplification of pre-existing health inequalities [1-3], both overall [4, 5], and in sub-population specific groups [6] (e.g., among Black, Asian and minority ethnic (BAME) groups, those experiencing socioeconomic disadvantage, and the unemployed). Evidence also suggests a worsening of mental health in adults [7, 8] and children [9], along with increases in loneliness and isolation [10-12]. Broader impacts on family functioning [13], loss of economic productivity [14], and education [15], including the gendered burden of home schooling have also been reported [16].

In contrast to the emerging evidence of the unintended negative effects of a national lockdown, here we report emerging evidence concerning unintended positive changes. Taking a salutogenic approach which focuses on the factors that support and promote health during stressful conditions [17], we examine the positive adaptation and growth experienced by some individuals. Using cross-sectional data from an online survey, the key objective of the current study is to examine the social patterning of the positive effects of lockdown across a range of domains. Given the large corpus of work concerning the role of structural social factors such as poverty, racism, gender and age in explaining health and illness we hypothesised that our findings would be shaped by these classic determinants of health.

## Method

### Participants and procedure

Data collection took place for 23 days from 20^th^ May 2020 to 12 June 2020, spanning the 9^th^ to the 12th week of national lockdown in Scotland. This national lockdown period was part of the UK-wide national lockdown that commenced in March 2020. The advice during national lockdown was to ‘stay at home’ and people were told to work from home wherever possible and to only leave their homes for essential purposes. These essential purposes included leaving home for food shopping, for medical purposes or to provide care for a vulnerable person. People were also allowed to leave home for one form of exercise each day. The target population of the survey was adults, aged 18 years or older, currently residing in Scotland, who were interested in sharing their experience of positive change. Participants were primarily recruited through social media advertisements on Facebook and Twitter which directed participants to the online survey on Qualtrics. All materials and procedures were approved by the Ethics Committee of the University of Strathclyde and all participants gave informed consent. The present sample comprised 3342 participants. Participant characteristics are shown in Table 1.

**Table 1.** Descriptive sample statistics of socio-demographic and health variables

| Variable |  | N | % |
| --- | --- | --- | --- |
| Household income | <£16 000 | 322 | 10.8 |
|  | £16 000-£29 999 | 591 | 19.9 |
|  | £30 000-£59 999 | 1185 | 40.0 |
|  | £60 000- £89 999 | 558 | 18.7 |
|  | £ 90 000+ | 316 | 10.6 |
|  | Missing | 370 |  |
| Employment status | Employed | 2258 | 67.6 |
|  | Inactive | 1033 | 31.0 |
|  | Unemployed | 48 | 1.4 |
|  | Missing | 3 |  |
| Highest education level | High School | 374 | 11.5 |
|  | College | 502 | 15.5 |
|  | Undergraduate | 988 | 30.4 |
|  | Postgraduate | 1380 | 42.6 |
|  | Missing | 97 |  |
| Age | 18-24 | 336 | 10.1 |
|  | 25-34 | 569 | 17.1 |
|  | 35-49 | 892 | 26.8 |
|  | 50-64 | 1130 | 33.9 |
|  | 65+ | 402 | 12.1 |
|  | Missing | 13 |  |
| Gender | Female | 2647 | 80.4 |
|  | Male | 646 | 19.6 |
|  | Missing | 49 |  |
| Ethnicity | White | 3215 | 97.0 |
|  | Non-White | 99 | 3.0 |
|  | Missing | 28 |  |
| Relationship status | Single | 602 | 18.2 |
|  | Married or living with partner | 2203 | 66.7 |
|  | Have a partner but not living together | 253 | 7.7 |
|  | Separated /divorced or widowed | 247 | 7.4 |
|  | Missing | 37 |  |
| Health | Very poor | 10 | 0.3 |
|  | Poor | 63 | 1.9 |
|  | Fair | 479 | 14.4 |
|  | Good | 1541 | 46.2 |
|  | Very good | 1243 | 37.3 |
|  | Missing | 6 |  |
| High-risk | Yes | 499 | 14.9 |
|  | No | 2842 | 85.1 |
|  | Missing | 1 |  |
| COVID-19 Diagnosis | Yes or suspected | 400 | 12.0 |
|  | Don't know | 978 | 29.3 |
|  | No | 1963 | 58.8 |
|  | Missing | 1 |  |

## Measures

### Sociodemographic and Health Variables

Participants provided sociodemographic and health data on the following variables: (1) gender, (2) age (18-24, 25-34, 35-49, 50-64, 65+), (3) relationship status (single, married/living with partner, living apart from partner, separated/divorced/widowed), (4) ethnicity, (5) education (high school, college, undergraduate, postgraduate), (6) annual household income (<£16,000, 16,000 – 29,999, 30,000 – 59,999, 60,000 – 89,999, £90,000+) (7) employment status (employed, inactive, unemployed), (8) overall health (very poor, poor, fair, good, very good), (9) risk status for COVID-19, e.g., aged 70+ or have an underlying health condition (yes, no), and (10) COVID-19 diagnosis (yes diagnosed, suspected, don’t know, no).

### Positive Changes

We measured positive changes using an expanded version of the positive events subscale of the Epidemic-Pandemic Impacts Inventory (EPII) [18]. We utilised 21 items to assess positive changes that participants may have experienced across a number of domains (e.g., relationships, physical activity, sleep, work). The measure that we utilised can be found via the project’s Open Science Framework page https://osf.io/nwh48/. Participants were asked “Since social distancing restrictions were introduced, what has changed for you?” with the response options of “yes”, “no” or “not applicable” across each domain. Example items included “more appreciative of things usually taken for granted”, “improved relationships with family and friends” and “increase in exercise or physical activity”. A positive score for each participant was computed by scoring 1 for a ‘yes’ and zero for ‘no’ on the 21 questions. A total score was computed which comprised of the sum of the ‘yes’ responses for each participant. This was then converted to a percentage score by dividing the total score by the sum of ‘yes’ and ‘no’ responses. NAs were removed from the analysis since individuals could not increase their score by responding to a question which did not apply to them. A higher percentage positive score indicated a greater proportion of positive changes. The scale was found to have good internal consistency (Cronbach’s α=.75) in the current study.

### Statistical analysis

Exploratory univariate analysis, taking each socio-demographic factor in turn, was performed to determine which factors were associated with the positive change scores. Between group scores were analysed using t-tests or ANOVA. Factors independently associated with positive change were then determined using a multiple regression model with stepwise variable selection in which variables are sequentially entered into the model. All analyses were done using Minitab (version 18) at a 5% significance level (adjusted for multiple comparisons were appropriate using the Tukey method).

## Results

### Descriptive statistics

The sample characteristics are shown in Table 1. In addition, a graph of the distribution of positive change scores is shown in Fig 2. Scores ranged from 0-100 with a mean of 47.2% (SD=20.8). Table 2 presents the proportions of people reporting yes, no, or not applicable at the item level of the positive change measure. From this it can be seen that the positive changes that were most commonly reported were: being more appreciative of things usually taken for granted (82.6%), more time doing enjoyable things (67.4%), more time in nature or being outdoors (65.3%), paid more attention to personal health (61.7%), increase in exercise or physical activity (53.9%), and more quality time with partner or spouse (53.3%).

**Table 2.** Item level proportions for the positive change measure

| **Item** | **Yes (%)** | **No (%)** | **NA (%)** |
| --- | --- | --- | --- |
| More quality time with partner or spouse | 53.3 | 21.2 | 25.4 |
| More quality time with children | 30.3 | 20.0 | 49.6 |
| Improved relationships with family or friends | 45.2 | 48.0 | 6.8 |
| New connections made with supportive people | 27.3 | 61.3 | 11.4 |
| Increase in exercise or physical activity | 53.9 | 45.1 | 1.0 |
| Increase in exercise or physical activity with family or partner | 41.9 | 47.4 | 10.7 |
| Discovery of new enjoyable ways to be physically active | 45.1 | 53.2 | 1.7 |
| Better quality of sleep | 33.3 | 64.1 | 2.6 |
| More time in nature or being outdoors | 65.3 | 33.7 | 1.0 |
| More time doing enjoyable activities | 67.4 | 31.6 | 1.0 |
| Developed new hobbies or activities | 41.9 | 57.1 | 1.1 |
| More appreciative of things usually taken for granted | 82.6 | 16.3 | 1.0 |
| Paid more attention to personal health | 61.7 | 36.9 | 1.4 |
| Paid more attention to preventing physical injuries | 40.9 | 55.0 | 4.0 |
| Ate healthier foods | 44.6 | 52.4 | 3.0 |
| Less use of alcohol | 23.1 | 59.6 | 17.4 |
| Spent less time on screens or devices outside of work hours | 12.6 | 83.8 | 3.6 |
| Volunteered to help people in need | 36.1 | 58.8 | 5.1 |
| Donated time or goods to a cause related to Covid-19 | 41.6 | 55.6 | 2.9 |
| Found greater meaning in work, employment, or studies | 30.2 | 58.3 | 11.5 |

**Fig 2.**
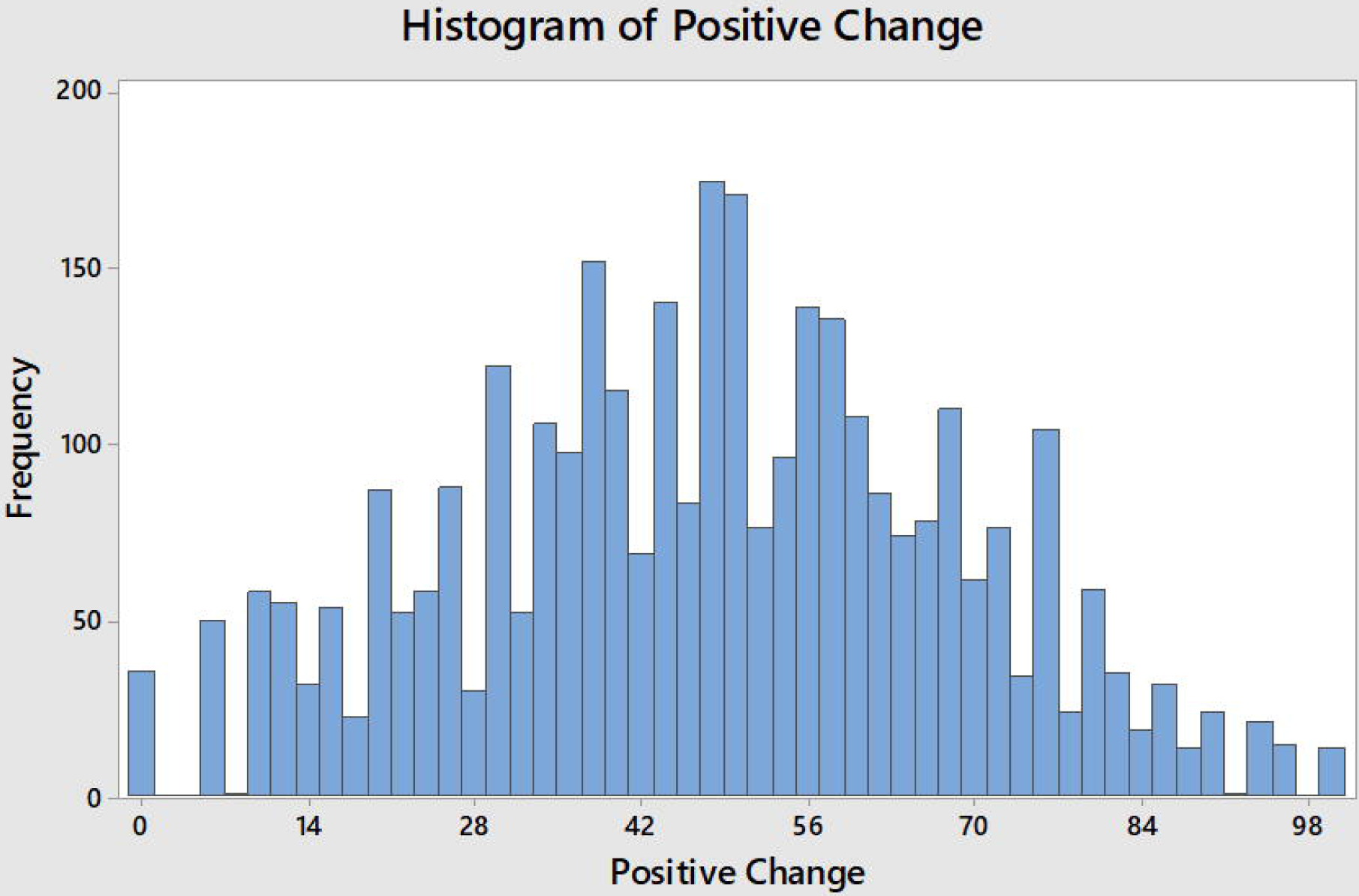
Distribution of positive change scores (%) in the sample.

### Positive Change Analysis

Univariate analyses were used to identify factors significantly associated with positive change with post-ANOVA comparisons where appropriate. Table 3 shows the results. Females reported significantly higher levels of positive change than males. In relation to age, we found that there was evidence of a difference in the mean positive change score between the age categories, with the older age group (65+) demonstrating the lowest level of positive change, and significantly lower than those aged 18-24. Similarly, there was evidence of significant differences in the mean scores across each of the relationship status groups, with those married or living with their partner exhibiting the highest level of positive change. Those in the employed group had higher levels of positive change than those in the inactive and unemployed groups (although the comparison with unemployed was not statistically significant due to the small number in the sample). When considering ethnicity, there was no evidence of a difference in positive change scores when comparing white and non-white participants. Similarly, for education, there was no evidence of a difference between the groups in terms of their positive change scores. For household income, there was no significant differences in positive change scores across the groups. However, there is a notable trend, with positive change score increasing with increasing household income levels.

**Table 3.** Univariate analysis of socio-demographic factors and positive change

| Variable | p-value | Comparison | Beta Coefficient | p-value |
| --- | --- | --- | --- | --- |
| Household income | 0.492 | £16-29.9K vs. <£16K | 0.64 |  |
|  |  | £30-59.9K vs. <£16K | 1.33 |  |
|  |  | £60-89.9K vs. <£16K | 1.84 |  |
|  |  | >£90K vs. <£16K | 2.54 |  |
| Employment status | 0.018 | inactive vs. employed | -1.92 | 0.014 |
|  |  | unemployed vs. employed | -4.89 | 0.107 |
| Highest education | 0.088 | college vs. high school | 1.19 |  |
|  |  | undergrad vs. high school | 0.37 |  |
|  |  | postgrad vs. high school | -1.25 |  |
| Age | 0.022 | 24-34 vs. 18-24 | -1.45 | 0.311 |
|  |  | 35-49 vs. 18-24 | -1.42 | 0.285 |
|  |  | 50-64 vs. 18-24 | -0.96 | 0.459 |
|  |  | >65 vs. 18-24 | -4.61 | 0.003 |
| Gender | <0.001 | male vs. female | -6.71 |  |
| Ethnicity | 0.423 | non-white vs. white | 1.70 |  |
| Relationship status | 0.010 | married/partner vs. single | 2.32 | 0.015 |
|  |  | partner (live apart) vs. single | -1.32 | 0.397 |
|  |  | sep/div/wid vs. single | 1.16 | 0.459 |
| Health | <0.001 | poor vs. very poor | 26.81 | <0.001 |
|  |  | fair vs. very poor | 27.16 | <0.001 |
|  |  | good vs. very poor | 29.99 | <0.001 |
|  |  | very good vs. very poor | 31.30 | <0.001 |
| Diagnosed | 0.390 | don't know vs. yes | -1.61 |  |
|  |  | no vs. yes | -1.46 |  |
| High risk | 0.008 | no vs. yes | 2.7 |  |

In relation to health, there was a significant effect of self-reported health, with those who reported their health to be ‘very poor’ having the lowest level of positive change and significantly lower compared to each of the other groups (poor, fair, good, very good). In addition, those that reported being at higher-risk of contracting COVID-19 had a significantly lower positive change score than those not at high-risk. Finally, there was no evidence of any difference in positive change scores based on COVID-19 diagnosis.

Multivariate analysis utilising stepwise regression showed that age, gender, relationship status, and self-reported health were all significantly associated with positive change. When considering the coefficients (see Table 4), it is shown that males reported a positive change score that was 6.1% lower than females. The older age group (65+) had a positive change score that was 7.5% lower than the younger age group. In terms of relationship status, those who were married or living with a partner had a score than was 3% higher than those who were single. When examining self-reported health, those who were in very poor health had a positive change score that was 24.5% lower than those in poor health, and 29.4% lower than those in very good health.

**Table 4.** Multivariate analysis of socio-demographic factors and positive change

| Variable | p-value | Comparison | Beta Coefficient | 95% CI | p-value |
| --- | --- | --- | --- | --- | --- |
| Age | <0.001 | 24-34 vs. 18-24 | -4.16 | -7.10, -1.22 | 0.006 |
|  |  | 35-49 vs. 18-24 | -4.69 | -7.53, -1.85 | 0.001 |
|  |  | 50-64 vs. 18-24 | -4.21 | -7.03, -1.39 | 0.003 |
|  |  | >65 vs. 18-24 | -7.54 | -10.85, -4.23 | <0.001 |
| Gender | <0.001 | male vs. female | -6.07 | -7.86, -4.28 |  |
| Relationship status | 0.001 | married/partner vs. single | 3.05 | 1.05, 5.05 | 0.003 |
|  |  | partner (live apart) vs. single | -1.76 | -4.82, 1.30 | 0.259 |
|  |  | sep/div/wid vs. single | 3.06 | -0.23, 6.35 | 0.068 |
| Health | <0.001 | poor vs. very poor | 24.49 | 9.30, 39.68 | 0.002 |
|  |  | fair vs. very poor | 25.79 | 11.48, 40.10 | <0.001 |
|  |  | good vs. very poor | 28.03 | 13.80, 42.26 | <0.001 |
|  |  | very good vs. very poor | 29.43 | 15.20, 43.66 | <0.001 |

## Discussion

The present study is the first to explore the social patterning of positive changes experienced during COVID-19 national lockdown. Referring back to Fig 1, there is clear evidence that unintended positive change has taken place as a result of lockdown, at least for some groups of the population. The important role of time was highlighted in the positive changes that had been made by the majority of the sample. Lockdown seems to have afforded people with more time to spend on activities they value. For example, the majority of the sample reported that they had been able to spend more quality time with their partner. In addition, participants reported that they had been able to spend more time doing enjoyable things, spend more time in nature or the outdoors, and increase their physical activity. Lockdown also seems to have provided participants with the time to reflect and the majority of participants reported that they were now more appreciative of things usually taken for granted. However, we found evidence of differences in the amount of positive change people had experienced, based on sociodemographic and health variables. Those groups with higher levels of positive change were females, those from younger age groups, people who were married or living with their partner, those who were employed, and those reporting better health.

The phrase ‘we are all in this together’ has been used, both domestically and internationally, throughout the pandemic to highlight the sense that COVID-19 is uniting us in shared experiences. However, there is a growing evidence base on the inequalities associated with COVID-19. Our findings fit within this emerging literature and point to the fact that while some groups were able to take advantage of lockdown as an unexpected opportunity to make positive changes in their lives, other groups were not. Similar findings on the inequalities associated with adverse experiences during lockdown have been reported [3], with the experience of more adverse events being related to socioeconomic position (consisting of household income, education, employment status, and housing). In addition, research on the experience of adverse mental health during COVID-19 has also shown the frequency of abuse, self-harm and thoughts of suicide/self-harm to be higher among women, BAME groups, those who were unemployed and those in poorer physical health [5]. Complementing this work, we also find that the experience of making positive changes in lockdown is shaped by many of these key sociodemographic factors.

Together these findings indicate the enduring nature of health inequalities and evoke key concepts from complex adaptive systems perspectives within public health [19, 20]. Despite the enormity of structural and social change that the national lockdown brought, there appears to be no sense of reaching a ‘tipping point’ in which the self-organising system that drives inequalities was radically disrupted or dramatically changed. In fact, emerging evidence suggests the opposite, there is clear evidence of negative feedback loops ensuring the system returned to stasis, reiterating inequalities along very familiar lines across a broad range of outcomes, for example, COVID-19 related morbidity and mortality, in addition to positive and negative psycho-social change.

We believe that our study is the first to report on the social patterning of positive changes during a period of COVID-19 national lockdown. The study also has the strength of a large sample size, and the inclusion of a wide range of sociodemographic factors. However, there are limitations. Most notably, as our participants were primarily recruited from social media our sample is not nationally representative of the Scottish population. In particular, in comparison to Scottish census data it is clear that we have an over-representation of female participants and those educated to University level. However, the sample has a good age distribution and the ethnicity and household income levels of the sample is broadly reflective of the Scottish population. A further consideration relating to recruitment via social media is that we may have reached a different type of participant than if we had been able to employ more traditional recruitment methods. However, as the study was conducted during national lockdown, we were restricted in the recruitment methods that were available. In addition, the sampling method employed was purposive. Participants responded to study adverts which asked them to share the positive changes they had made during lockdown. We used this sampling strategy deliberately as we wanted to recruit participants who had experienced positive change in order for us to examine the processes behind these positive experiences. However, the amount of positive change being reported by participants in our study may not be typical of the experiences of the general population. In addition, the study is cross-sectional in nature and so provides only a snapshot of the positive changes people were experiencing at a particular stage of the lockdown, and cannot, at this stage, provide data on whether these positive changes were maintained over time.

It is also important to consider the that the positive changes experienced by participants within the context of a national lockdown in Scotland may be different from the positive changes that people living in other national lockdown contexts experienced. As noted above, time was central to many of the positive changes that people made. National lockdowns across the board are likely to have afforded many people more time as time commitments such as commuting and many forms of socialising were removed. This extra time is likely to have provided many people with an opportunity to reflect, and as noted by the participants in our study to be more appreciative of things usually taken for granted. In this regard, our findings are likely to be applicable to other national lockdown contexts. However, they may differ from national lockdown contexts where stricter restrictions were imposed on time outdoors and time allowed for exercise. Within the national lockdown in Scotland people were allowed to leave their homes for exercise and many of our participants noted that they had been able to spend more time in nature and had increased their physical activity levels. Indeed, recent research has reported that moderate-to-vigorous physical activity levels increased during the national lockdown in Scotland [21]. These types of positive change would not have been possible in countries with stricter national lockdowns where people were not allowed to leave their homes for exercise.

## Conclusion

The present study reports preliminary evidence relating to the social patterning of self-reported positive change during lockdown. The data reported here are part of the larger mixed-methods CATALYST project which seeks to understand how people have initiated and maintained positive change across a number of domains, during lockdown, and as restrictions have been eased. The aim is to share this learning through intervention development work, in order to facilitate positive health change in others. From the results of the current study, we can see that there are sub-populations and communities where it may be particularly important to target these interventions, in order to provide opportunities for health change amongst those who may benefit most.

## Data Availability

The data will be available on publication of the paper on the Open Science Framework http://doi.org/10.17605/OSF.IO/NWH48

## Acknowledgements

The study was funded by the Chief Scientist Office, Scottish Government (Ref: COV/SCL/20/09).

